# Time-varying effects are common in genetic control of gestational duration

**DOI:** 10.1101/2023.02.07.23285609

**Authors:** Julius Juodakis, Karin Ytterberg, Christopher Flatley, Pol Sole-Navais, Bo Jacobsson

## Abstract

Preterm birth is a major burden to neonatal health worldwide, determined in part by genetics. Recently, studies discovered several genes associated with this trait or its continuous equivalent – gestational duration. However, their effect timing, and thus clinical importance, is still unclear. Here, we use genotyping data of 31,000 births from the Norwegian Mother, Father and Child cohort (MoBa) to investigate different models of the genetic pregnancy “clock”. We conduct genome-wide association studies using gestational duration or preterm birth, replicating known maternal associations and finding one new foetal variant. We illustrate how the interpretation of these results is complicated by the loss of power when dichotomizing. Using flexible survival models, we resolve this complexity and find that many of the known loci have time-varying effects, often stronger early in pregnancy. The overall polygenic control of birth timing appears to be shared in the term and preterm, but not very preterm periods, and exploratory results suggest involvement of the major histocompatibility complex genes in the latter. These findings show that the known gestational duration loci are clinically relevant, and should help design further experimental studies.

## Introduction

Preterm birth – birth before completing 37 weeks of gestation – is a major cause of neonatal mortality and morbidity, affecting about 10 % of deliveries worldwide (Vogel et al., 2018). Various environmental factors associated with preterm birth have been reported, but the biological mechanisms leading to it are still largely unknown. As a result, there is no robust strategy for prevention of preterm birth, and its rates continue to increase in many countries (Blencowe et al., 2012; Jing et al., 2020; Vogel et al., 2018).

Genetics is clearly important for this trait: around 20–40 % of the variation in pregnancy duration is attributed to maternal genetic factors, and another 10–30 % to foetal genetics (Wadon et al., 2020). Although the specific variants or genes underlying these effects are still mostly unknown, several loci were recently identified in large genome-wide association studies (Liu et al., 2019; Solé-Navais et al., 2022; Zhang et al., 2017). Understanding the biological effects of variants at these loci could lead to broader understanding of pregnancy maintenance, and suggest possible interventions for preterm birth.

Various statistical models have been used for discovering the variants, and their comparison could provide some clues about the mechanism of the pregnancy “clock”. Typically, the variants are tested for association with pregnancy (gestational) duration in days as a continuous outcome using linear models, or preterm birth as a binary outcome, using e.g. logistic regression. Other dichotomizations of pregnancy duration have also been used, such as early (Liu et al., 2019), extreme (Rappoport et al., 2018) or very preterm birth (Tiensuu et al., 2019), or post-term birth (Liu et al., 2019). So far, almost all of the associated variants have been identified using the continuous outcome, and have weaker or no associations with preterm birth (Liu et al., 2019; Solé-Navais et al., 2022; Zhang et al., 2017).

This contrast between results for pregnancy duration and preterm birth raises questions about the action and clinical relevance of the discovered genes. On one hand, if their variants have uniform effects throughout pregnancy (as modelled in the standard linear regression), such results are expected, as the continuous model then has much higher detection power (Ragland, 1992). On the other hand, pathways initiating preterm birth are considered to be distinct from those acting near term (Romero et al., 2014). This is supported by epidemiological associations that differ by gestational duration groups (Auger et al., 2011), and registry data also suggests that genetic effects change throughout pregnancy (Juodakis et al., 2017). It is possible that the loci discovered for gestational duration have little relevance for preterm birth – or they might act uniformly throughout pregnancy and contradict this view.

In this study, we aim to resolve this uncertainty. We analyse a genotyping cohort of more than 30,000 parent-offspring trios using models explicitly incorporating the timing of genetic effects. With these, we aim to answer: 1) are the pregnancy duration genes discovered so far clinically relevant for preterm birth; 2) do the genes support the view that preterm and term birth timing is aetiologically different; 3) is there a period of gestation where the gene effects are particularly strong, weak or otherwise unusual.

We start by conducting genome-wide association studies (GWASs) of gestational duration and preterm birth in this cohort, and directly show the challenges in comparing the results of the two outcomes. We then select the top variants known for these traits and re-analyse them in time-to-event models, specifically designed to investigate their effect dynamics. We also repeat this analysis on a polygenic score of gestational duration, and further explore possibly different mechanisms acting in the very preterm range.

## Results

### GWAS results for gestational duration and preterm delivery

We ran maternal and foetal GWASs of gestational duration and preterm delivery on 22,247 mothers and 21,262 foetuses (610 and 529 preterm cases, respectively) from the Norwegian MoBa cohort, using standard association methods (linear and logistic regression). In the maternal GWAS of gestational duration, we identified two loci associated with gestational duration at genome-wide significance (**Figure 1A**), located in or near the *KCNAB1* and *ADCY5* genes. Both of these loci have been reported previously in (Solé-Navais et al., 2022), however *KCNAB1* was not identified in another large GWAS (Zhang et al., 2017). In the foetal GWAS of gestational duration, one single genome-wide significant variant was identified, rs34207099, nearest to the gene *LRATD2* (*FAM84B*) (beta = −2.4 days, SE = 0.44 days, p = 3.0e-8, imputation INFO score = 0.75, **Figure 1C**). We note however that the variant has a low frequency (MAF = 1.1%), is close to the significance threshold, has no linked proxies, and this association is not observed in the previous, larger foetal meta analysis (p = 0.20) (Liu et al., 2019). For preterm delivery, no genome-wide significant associations were detected either in the maternal (**Figure 1B**) or foetal GWAS (**Figure S1**). Genomic inflation was small in all four GWASs, with lambda <1.06 (**Figure S2**).

**Figure 1.**
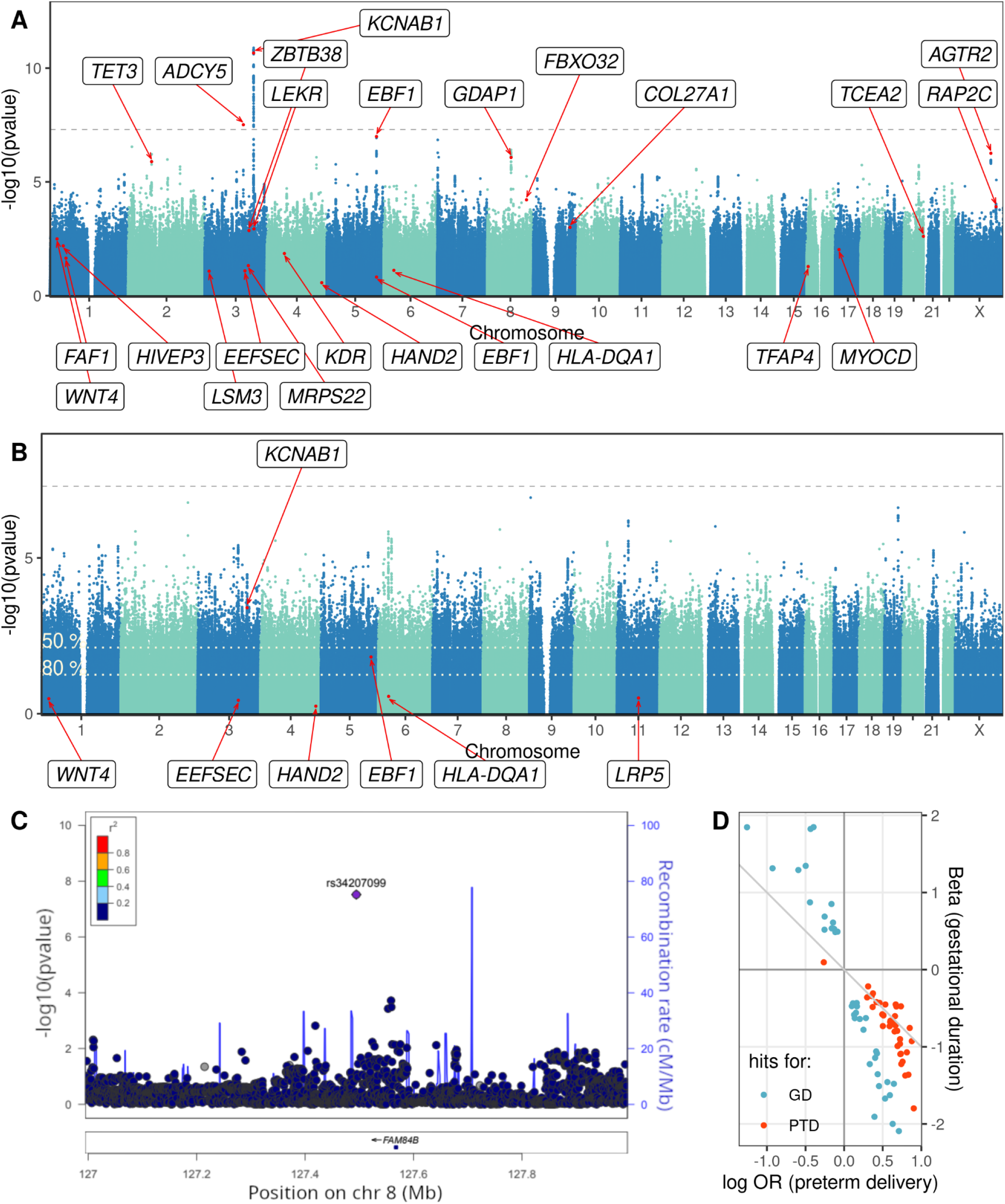
(A,B) Manhattan plots illustrating the maternal GWAS results for gestational duration (A) and preterm delivery (B). The dashed line marks the significance threshold. The variants marked in red, and the corresponding loci, were reported as significant for the respective phenotype in the largest previous GWAS meta-analysis (Solé-Navais et al., 2022). Under the liability model, the GWAS in (B) has lower power: for a variant that can be detected genome-wide with 80 % power in (A), the significance thresholds in (B) should be reduced to the dotted white lines to achieve 50 % or 80 % detection power. (C) LocusZoom plot showing the region around the only genome-wide significant association identified in the foetal GWAS. Shown p-values are for gestational duration. The significant variant is the purple diamond while all other variants are coloured based on their linkage disequilibrium with it. Variants not available in the reference panel are coloured in grey. (D) Effect size comparison for the suggestive variants in the maternal GWASs (p-value < 10^−5^ for either outcome). Shown are the log odds ratios (OR) and linear regression betas in the same position. Identity line is marked.

The null results of the preterm delivery GWAS at first glance suggest differences from the continuous phenotype. Of the 165 preterm delivery loci at suggestive association (p < 10^−5^) none were (genome-wide) significant in the GWAS of gestational duration. However, the regression coefficients of the suggestive variants across the two maternal GWASs were concordant in direction and size, i.e. variants that shorten gestational duration also increase preterm delivery risk (**Figure 1D**). If the genetic effects are indeed shared between term and preterm periods, then dichotomization incurs a great loss of power: we estimate that such analysis would then require a 6.7x larger cohort to reach equal power with the continuous analysis (see also **Figure 1B;** calculations based on 2.7 % prevalence and the liability model, see Methods). In light of this power difference, comparison between the two GWAS is complicated, and we could not make definite conclusions about the effect dynamics or clinical relevance of the findings.

### Top gestational duration variants have time-varying effects

From previous meta-analyses, we identified 25 maternal and 3 foetal genetic variants involved in delivery timing. One additional foetal locus was identified in the present study. Most (26) of these were initially discovered in analyses of gestational duration, although one (maternal) variant was detected only for preterm delivery and two (foetal) only for early preterm birth.

We then re-analysed these 29 variants with a flexible survival model (PAMM), and found that many of them have time-varying effects. PAMM (see Methods for detailed presentation) explicitly models the time to event, i.e. delivery. The effect of a variant is the change in instantaneous risk to deliver, relative to the major allele, and is allowed to change during gestation. **Figure 2** shows the estimated effect dynamics for 20 variants that were nominally associated with gestational duration in this cohort, and 14 of these were significantly changing over time (p < 0.05 for the non-linear term in PAMM). Notably, even though all of the 20 variants were discovered in linear models and cohorts with few preterm births, their effects generally do not appear to be limited to the term period. In fact, for 10 loci (*KCNAB1, FBXO32, LRATD2, ADCY5* and others) the estimated effect was largest early in the pregnancy, and decreased towards term. In three other cases, the effect peaked in the preterm birth range, around week 34 (*TET3, EBF1, COL27A1*). For the remaining loci, the confidence intervals are wide and we are cautious about interpreting the estimated shapes, but most still show effects around or before 37 weeks (**Figure 2**). One exception is the *IL1A* locus, which only had a significantly non-zero effect during weeks 38 to 41, consistently with the analyses performed in (Liu et al., 2019), which also concluded that its effects are mostly seen around term.

**Figure 2.**
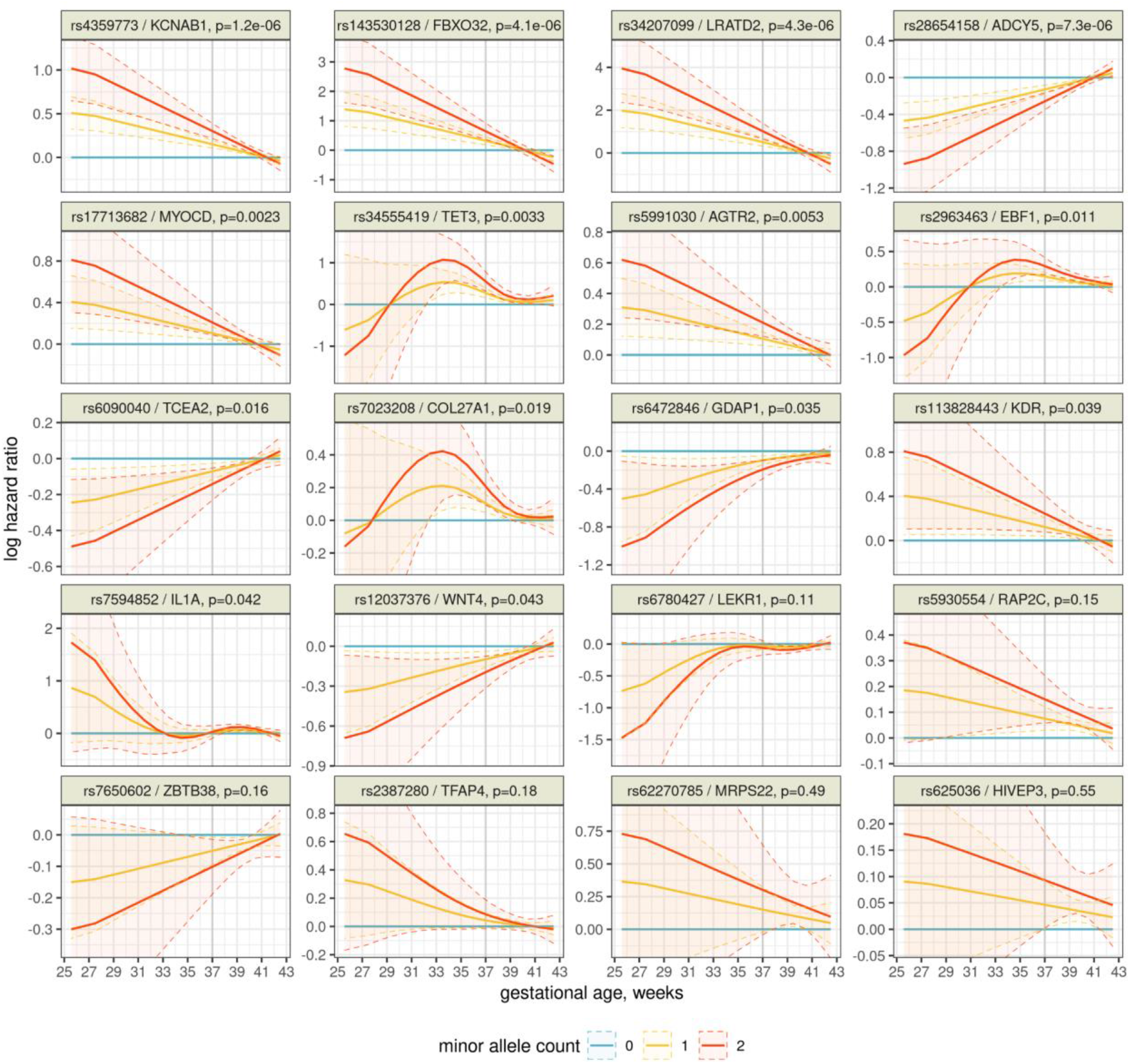
Effects of 20 genetic variants on delivery timing throughout gestation, estimated by a flexible PAMM. For each week of gestation, the line is the estimated instantaneous log hazard ratio (positive = risk of birth is currently increased) and its 95 % confidence interval is shaded. The preterm limit of 37 weeks is highlighted. Note that the minor allele count was used as a continuous covariate in the analysis, so the effects of 1 and 2 minor alleles are constrained to be proportional.

The remaining 9 out of 29 variants are shown in **Figure S3**. Eight of these variants did not have any detectable effects on delivery timing in this cohort (p>0.05 both in the gestational duration GWAS, and for the non-linear effects in PAMM). This includes all three loci discovered in dichotomized analyses (*LRP5, SPATA6, LPP*). The *EEFSEC* locus had borderline associations (p = 0.06 for the linear effects in GWAS, p = 0.02 for the non-linear effects in PAMM) and is also shown in **Figure S3**. Details of all 29 variants are listed in **Table S1**.

As diagnostics of our PAMM setup, we verified that it maintains the type I error rate in this setting by simulations (p < 0.05 observed in 4 % of simulations), and that if only constant effects are included, the estimates are very similar between PAMM and standard Cox regression (**Figure S4**).

As a sensitivity analysis, we applied two other methods for testing time-varying effects. These methods assume different (linear) shapes of the effects, and produce larger p-values for the non-linear effect loci (**Table S1**). The ranking of most other loci, and the overall number of significant results, are similar across the methods.

We also confirmed that almost all of the 28 top loci genes were actively transcribed in the placenta during early gestation. (Note that there were 29 loci, but two map to the *EBF1* gene.) In the data from (Breen et al., 2020), only *AGTR2* and *LEKR1* had few detections, while all other genes had at least 5 counts in 95 % of the samples. Overall, the expression level of our top loci genes (median count = 326, taken over samples and then over genes) was at least as high as of other protein coding genes (median count = 139; see also **Figure S5**). This shows that the genes discovered in term-centric analyses are already present in the transcriptome, and could plausibly have effects, much earlier in gestation.

### Polygenic effects are shared at term and early, but not very early gestation

We constructed a maternal polygenic score (PGS) for gestational duration based on the summary statistics of a previous large meta-analysis. The score includes ∼1.1 million variants, and on its own explains 2.0 % of the variance of gestational duration in the present cohort (or 2.8 % in combination with the clinical covariates). For comparison, if this score is limited to 50 kbp regions surrounding the 25 top hits, the variance explained drops to 0.6 %, or 1.5 % with clinical covariates.

Firstly, we confirmed that the estimated time-varying effects of each variant remain after adjusting for this score; they were also robust to adding or removing the clinical covariates (**Figure S6**).

We then analysed the effects of this score in time-to-event models (**Figure 3A**). Similar to the individual variants, the PGS shows effects extending well into the preterm range, even though it was derived based on analyses of continuous gestational age. Surprisingly, the “protective” PGS groups appear to have neutral or even slightly harmful effects in the very early gestation. While there is large uncertainty in the estimates, the pattern is still seen with different groupings of the score (**Figure S7**), or even by just directly observing the risks of different outcomes in each group (**Figure 3B**): the risk for preterm delivery decreases smoothly with the PGS, but for delivery before 32 weeks (or very preterm delivery, vPTD) the risk is similar across four PGS groups and only elevated in one.

**Figure 3.**
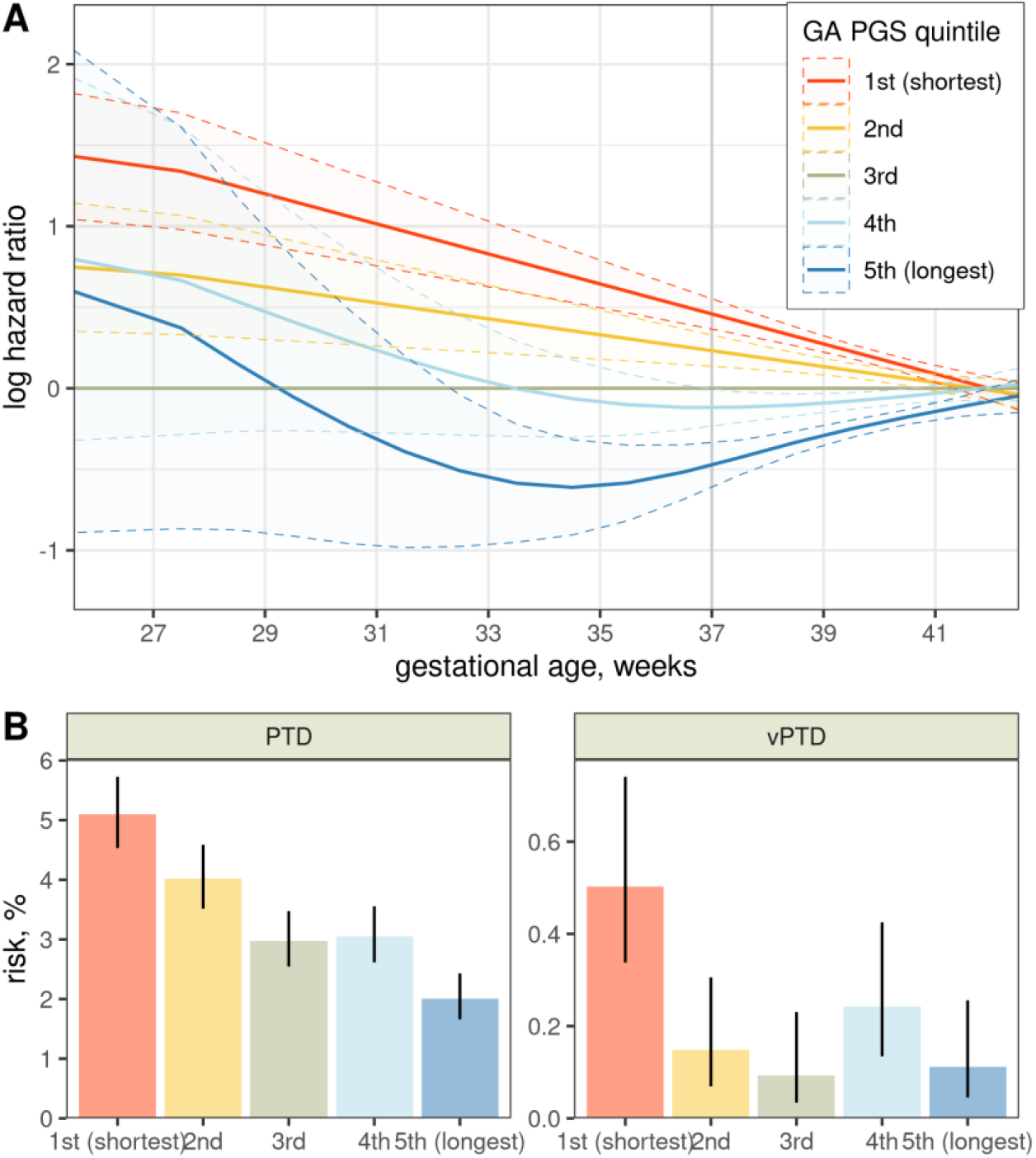
(A) Effects of the gestational duration polygenic score throughout gestation, estimated by a flexible PAMM. For each week of gestation, the line is the estimated instantaneous log hazard ratio (positive=risk of birth is currently increased), and its 95 % confidence interval is shaded. (B) Unadjusted risks of preterm (left) and very preterm (right) delivery in each polygenic score quintile. Lines show the 95 % confidence intervals.

We tested some possible explanations of this pattern (although we caution that the data contains only 59 vPTD cases). For example, it is biologically plausible that vPTD risk is determined mostly by rare alleles with strong deleterious effects; but the frequency of such alleles should decrease smoothly across all PGS groups, and indeed we observed that for rare deleterious alleles in the PGS (**Figure 4A**). Similarly, the number of homozygous deleterious genotypes decreased smoothly with PGS, not enriched in any particular group (**Figure 4A**). Neither of these counts was predictive of vPTD (i.e. no association after adjusting for PGS, p = 0.37 and 0.41). We then tested the top principal components, seeking to identify any broader genetic features related to vPTD, and found that only the third PC was significantly associated with it (p = 0.016). Also, this PC was not associated with all PTD (p = 0.16). Inspecting it revealed that its loadings were higher in chromosome 6, and specifically in the major histocompatibility complex region, MHC, 6p22.1–6p21.3 (**Figure 4B**). This suggests that immune pathways may be particularly relevant to the genetic risk of vPTD, possibly through gene interactions or other complex mechanisms producing the observed non-linear relation with PGS.

**Figure 4.**
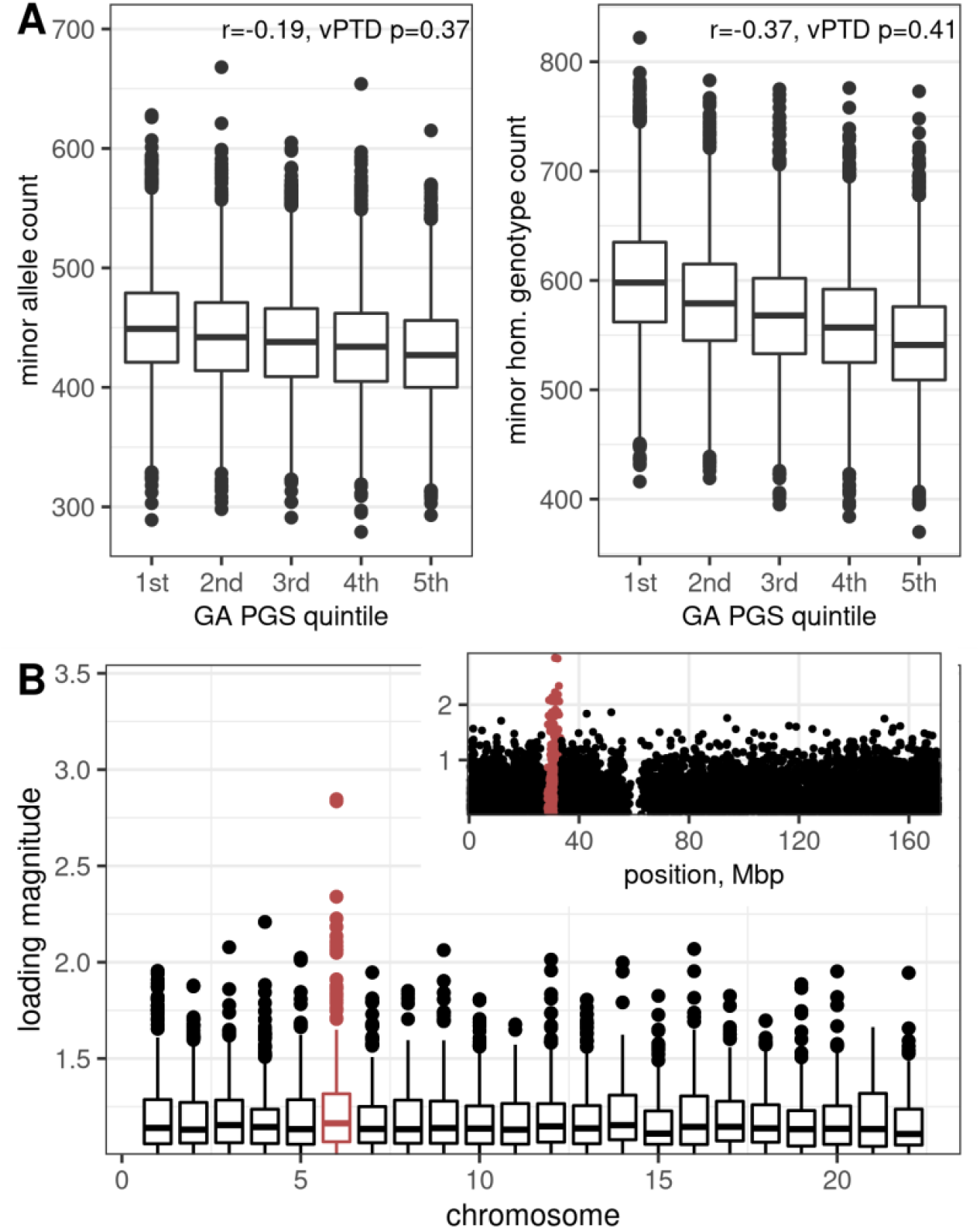
(A) Boxplots showing counts of rare alleles (left) or minor homozygous genotypes (right) across groups of gestational duration PGS. Only variants with estimated negative effects are shown here. Also shown are the correlation *r* between the count and the PGS, and p-value for the count term in a model of vPTD. (B) Variant loadings of the vPTD-associated principal component 3. Boxplots show the loadings across chromosomes, excluding any with magnitudes smaller than 1. Chromosome 6 is highlighted, and the inset shows all its loadings. The MHC region 6p22.1–6p21.3 is highlighted in the inset.

## Discussion

In summary, we conducted GWASs of gestational duration, noting 3 associated loci, and of preterm birth, which did not find any significant associations. Together with existing genome-wide studies, this brings the total number of loci known to be involved in pregnancy length to 29. Re-analysing these loci, as well as a gestational duration PGS, in time-to-event models showed that time-varying effects are common, and are often strongest in the preterm period. The PGS results and exploratory analyses also suggest that very preterm birth may be controlled differently, possibly involving the major histocompatibility gene region.

Our maternal GWAS results, in terms of the top variants, are largely similar to previous ones obtained from a direct-to-consumer genotyping cohort (Zhang et al., 2017). This cohort substantially differed from ours in phenotype ascertainment, exclusion criteria, and preterm delivery rates (which in Norway are relatively low (Vogel et al., 2018)), so the agreement between results is encouraging for the generalizability of GWASs. The weak foetal genetic associations are also consistent with literature (Liu et al. 2019). Still, cohorts focusing on non-European ancestries, and especially high case rate populations, are largely missing, and will be crucial to further progress in the genetics of pregnancy.

Our primary finding of practical importance is that many of the variants discovered for gestational duration also have strong effects in the preterm period. In other words, the lack of associations with preterm delivery does not mean lack of effects – rather, the power loss when dichotomizing is just too great (this is difficult to quantify directly under time-varying effects, but we showed a large loss under the simple liability model). Researchers thus should be encouraged to use gestational duration as a continuous phenotype which leads to clinically relevant findings. Stronger early effects in survival models can also be a technical artefact of frailty – unaccounted heterogeneity between the individuals, typically due to genetics or other covariates (Balan & Putter, 2020). However, as we observed several different effect shapes across the loci, and the effects were unchanged after accounting for known clinical covariates or the genetic background (PGS), we do not believe that frailty was a major factor in this study. Alternatively, such data could be analysed with accelerated failure time models, which are not impacted by frailty (Crowther et al., 2022), but they still require some development of significance tests and computational tools before practical use.

We also find that, while the gene effects are often present at some level throughout pregnancy, their strength greatly varies over time. This suggests that the biological clock controlling pregnancy length does not “tick” evenly, but rather has distinct phases at which different genes become relevant. A similar theory has been proposed for miscarriage timing – namely that there are distinct checkpoint times at which foetal development is evaluated and the pregnancy continued or not (Baird, 2009). In relation to preterm birth, different, simpler, clock models have been discussed in (Rokas et al., 2020), but here we present the first data from human genomics directly showing the complexity of this regulation.

Further functional work will still be needed to clarify the mechanisms behind the associations observed here. In particular, it is possible that the relevant genes directly act very early in pregnancy, e.g. in the trophoblast invasion (Tiensuu et al., 2019), but the impact of that only becomes apparent later. This is compatible with the decreasing effects that we observe for many genes (Jiang et al., 2021). On the other hand, we do show that our loci are actively expressed in early-to-mid-gestation placenta, which supports a more persistent role throughout pregnancy, and is also notable as many transcripts are missing in the placenta (Gong et al., 2022). In any case, we expect that follow-up studies can use our results to prioritise the most relevant genes and periods for each gene.

Finally, the analysis framework presented here can be applied to other exposures and outcomes as well. Similar questions about the timing of effects have been investigated with other factors of preterm birth as well, but the studies so far used ad hoc techniques, such as separate logistic regression models within subgroups (Auger et al., 2011). Moreover, there is great interest in applying time-to-event models to the age of onset of various adulthood diseases (He & Kulminski, 2020; Jiang et al., 2021; Pedersen et al., 2022). In both cases, adopting the procedures presented here could provide easier interpretation, likely more precise effect estimates (due to the use of continuous data across the full range of the phenotype), and more direct testing of clinically and biologically relevant questions.

## Methods

### Data source

The Norwegian Mother, Father and Child Cohort Study (MoBa) is a population-based pregnancy cohort study conducted by the Norwegian Institute of Public Health (Magnus et al., 2016). Participants were recruited from all over Norway in 1999–2008. The women consented to participation in 41 % of the pregnancies. The cohort includes approximately 114,500 children, 95,200 mothers and 75,200 fathers. The data used in this study is version 12 of the quality-assured data files released by MoBa, and includes genotype data, questionnaires filled out by the parents, and linked records from Medical Birth Registry (MBRN), a national health registry containing information about all births in Norway.

The establishment of MoBa and initial data collection was based on a licence from the Norwegian Data Protection Agency and approval from The Regional Committees for Medical and Health Research Ethics. The MoBa cohort is currently regulated by the Norwegian Health Registry Act. The current study was approved by the Norwegian Regional Committee for Medical and Health Research Ethics South-East (2015/2425) and Swedish Ethical Review Authority (Dnr 2022-03248-01).

### Study population and genotyping

The present study uses a subset of this cohort: pregnancy outcomes, maternal and foetal genotypes from 31 496 parent-offspring trios that were genotyped over 2012–2018. We note that these samples have been used as a small part of a previous meta-analysis of gestational duration (Solé-Navais et al., 2022). Genotyping included only singleton, live-birth pregnancies with complete birth registry data and at least the first MoBa questionnaire answered, and individuals alive at time of genotyping. Further, we excluded pregnancies that used IVF, where the mother had diabetes, the child’s APGAR score was 0 at both 1 and 5 min, recorded gestational duration was an extreme outlier, and kept one random pregnancy for each mother that had repeated pregnancies in the cohort.

Genotyping was performed in nine batches on different Illumina arrays (HumanCoreExome-12 v1.1, HumanCoreExome-24 v1.0, Global Screening Array v1.0, InfiniumOmniExpress-24 v.2, HumanOmniExpress-24 v1.0). Genotypes were called in Illumina GenomeStudio software (v.2011.1 and v.2.0.3). Cluster positions were identified from samples with call rate ≥ 0.98 and GenCall score ≥ 0.15. Variants were excluded if they matched any of: GenomeStudio parameters of cluster separation < 0.4, 10 % GC-score < 0.3, AA T Dev > 0.025, or call rate < 98 % or HWE p-value < 1e-6. Samples were excluded if they showed < 98 % markers called, heterozygosity excess > 4 SD, genotyped sex mismatch, or were related to another sample in the cohort with estimated identity proportion (PI_HAT) > 0.1.

Finally, individuals of primarily non-European ancestry were excluded based on PCA with reference samples from the HapMap project, version 3.

Pre-phasing was conducted locally using Shapeit v2.790 (O’Connell et al., 2014). Imputation was performed at the Sanger Imputation Server with Positional Burrows-Wheeler Transform and HRC version 1.1 reference panel (Van den Berg et al., 2016). All coordinates are provided in reference to the human genome build GRCh37.

### Genome-wide association studies

Four genome-wide association studies (GWASs) were performed in this data, using maternal or foetal genomes and gestational duration (in days, determined by ultrasound) or preterm delivery (gestational duration < 259 days) as the outcome. The analyses were performed in PLINK v2.00 alpha 3.3 (Chang et al., 2015) using an additive regression model (linear or logistic) with minor allele dosage and batch covariates. Non-spontaneous deliveries (initiated by induction or caesarean) and genetic variants with minor allele frequency <1 % were excluded here. Standard genome-wide significance threshold of 5e-8 was used. Lambda and intercept parameters for genomic inflation were calculated using LD score regression (Bulik-Sullivan et al., 2015) and pre-computed LD scores obtained in subjects of recent European ancestry from the 1000 Genomes Project. The regional scatter plot for the foetal locus was done using the LocusZoom’s web application (Pruim et al., 2010). The LD reference panel was 1000 Genomes Mar 2012 EUR cohort.

For comparing the effect directions, variants at suggestive association in both of the maternal GWASs (p < 1e-5) were selected, and pruned by distance (i.e. removing other variants within 1.5 Mbp).

For comparing the GWAS power for the two phenotypes, we used formulas from (Wu & Sham, 2021). Specifically, assume that the gestational age distribution is normal, and variants only have additive, constant effects on its mean, not in any way specific to preterm or other time periods. This is the liability model, and then the sample size required to achieve equal power in continuous and dichotomized analysis (namely, Wald tests of simple linear or logistic regression coefficients) is related by a factor of *t*^2^, where 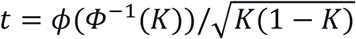, *K* is the prevalence of cases, and *ϕ, Φ*^−1^ – the pdf and quantile function of the standard normal distribution, respectively. The same factor relates the continuous and dichotomized effect sizes and SEs: *β*_*d*_*/SE*_*d*_ = *tβ*_*c*_*/SE*_*c*_. The significance level allowing power *f* in the dichotomous analysis is *Φ*(−*Φ*^−1^(1 − *f*) − *β*_*d*_*/SE*_*d*_); we calculated this for effect size corresponding to the minimum standardised effect detectable in the continuous analysis at genome-wide significance and 80 % power, i.e., *β*_*c*_*/SE*_*c*_ = *Φ*^−1^(0.8) − *Φ*^−1^(5 × 10^−8^/2), converted as above.

### Time-to-event analysis of top variants

For detailed analysis of timing, we selected all loci identified as genome-wide significant for pregnancy duration or preterm birth in the largest available maternal (Solé-Navais et al., 2022) and foetal (Liu et al., 2019) meta-analyses. (All loci discovered in the third large study by (Zhang et al., 2017) are represented by at least one variant in these results.) From each locus, the top variant as initially reported was chosen. One new foetal variant identified in the present study was added to this set. The name of each locus here is the primary gene assigned to it in the source study.

The variants were re-analysed in time-to-event models based on the Cox proportional hazards framework. (See (Kleinbaum & Klein, 2006) for a general introduction to such models, or (Juodakis et al., 2017) about their use with pregnancy duration.) Our event of interest is spontaneous delivery. The hazard *h*(*t*) then is defined as the instantaneous risk of delivery at time *t* for a mother who has not delivered before *t*. The covariates **x** are modelled to change this risk log-linearly:

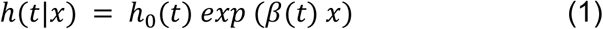

Where *h*_0_(*t*) is a baseline hazard, and *β*(*t*) are the effects of covariates. Note that this effect may vary in time, e.g. a covariate may be only relevant during some particular periods of pregnancy. Inspecting the shape of *β*(*t*) for each selected variant, and testing whether it is constant, are the main aims of our analysis.

Our primary approach for computing this uses an approximation of (1): the piecewise exponential additive mixed model, PAMM (Bender et al., 2018). Its idea is to partition the range of observed event times into short intervals, within which the hazard is assumed to be constant and the number of events is Poisson-distributed. Given sufficiently short intervals, the likelihood of this model is proportional to that of (1), so this way existing tools for Poisson regression and generalised additive mixed models can be used to fit model (1). In particular, the baseline hazard and the effect *β*(*t*) can be expanded using a spline basis to obtain smooth non-linear forms (Bender et al., 2018; Wood, 2017).

Specifically, for each variant, we fit the following hazard rate model:

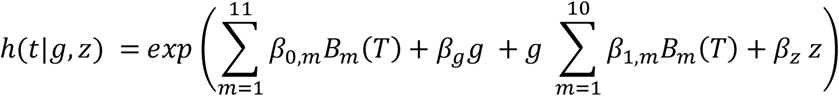

Where *T* is the end time of the interval containing *t, B*_*m*_ the basis functions of a cubic regression spline, *g* the minor allele dosage and other covariates (with constant effects) **z**. The model is fit using R packages *mgcv* (Wood, 2017) and *pammtools* (Bender et al., 2018). Our null hypothesis is that the variant has only constant effects, i.e. all *β*_1,*m*_ = 0. We test its significance and extract the estimated effects over time and their confidence intervals using standard *mgcv* functionality (Wood, 2017). We also checked that this test maintains the type I error rate in our setting: we simulated phenotype by bootstrap from the observed distribution in the sample, simulated a locus with no effect and minor allele frequency of 0.3, and fitted the model, in 2000 replicates.

The outcome and covariates were prepared as follows. The pregnancy durations were shifted to start at 1 (by subtracting 169 days), so that the period where no live deliveries are expected would not affect the estimation. Non-spontaneous deliveries were treated as censored. The PAMM interval endpoints were set to 0, 20 and then every 7 days. Minor (within this dataset) allele dosage was used as retrieved from the imputation, i.e. a number between 0–2, with occasional non-integer values representing uncertain imputation. The other covariates were genotyping batch, maternal age (in years) as a pair of degree-2 orthogonal polynomials, year of the delivery, maternal height (in cm), foetal sex (reference level male), presence of congenital malformations (reference level no), and parity (reference 1, other categories 0, 2, or ≥ 3 previous deliveries). For maternal height, missing or extreme outlier values were mean-imputed.

To verify that our PAMM setup approximates the Cox model (1), at least when the effects are constant, we ran a simplified PAMM,

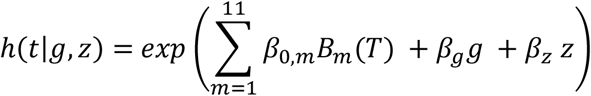

And a Cox regression with the same covariates, as implemented in R *survival* package (Therneau, 2022), and compared their *β*_*g*_ estimates. As a sensitivity analysis, two other tests for non-proportionality of hazards were applied. First, a PAMM that includes a *time x genetic effect* interaction as a specified form (linear), instead of the spline, i.e.:

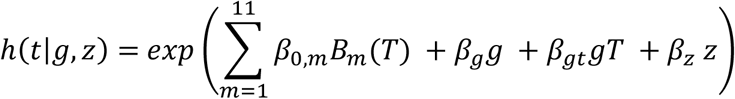

The significance of the interaction term, as calculated by *mgcv*, is reported. The second test was the cox.zph test implemented in the *survival* package; it again reports the significance of the interaction term, but here the times are replaced with the corresponding Kaplan-Meier survival estimates *S*(*t*) to reduce the effect of outliers, i.e. the term is *β*_*gt*_*g* (1 − *S*(*t*)) (Therneau, 2022).

As a functional support for the observed effects of the top loci, we also inspected the expression levels of the corresponding genes in RNA sequencing data from (Breen et al., 2020). The data was obtained from 125 placenta samples, collected at pregnancy termination between 6–23 weeks of gestation. The read counts for each gene were retrieved from GEO (GSE150830), and we removed non protein coding genes based on Ensembl annotations. We report the median of sample counts for each gene, and also the median of these over genes in each set (top loci or all other).

### Construction and analysis of the gestational duration polygenic score

We calculated a gestational duration polygenic score for each mother included in the study. The score weights were previously trained in a maternal GWAS meta-analysis on gestational duration (Solé-Navais et al., 2022). The MoBa cohort of the present study was excluded, resulting in n=173,162 mothers for the GWAS, and the score weights were trained using LDpred2. Each individual’s polygenic score is the product of the weighted effects of the variants from the training and the number of corresponding effect alleles. In this section, we derived hardcall genotypes for the study population, and calculated the individual scores using PLINK 1.9. Note that foetal scores were not used here due to low overall heritability. The phenotype was prepared as described in the previous sections, with induced births censored in the PAMMs and excluded in all other models.

The variance explained by the PGS was reported as the R-squared from a linear regression model of gestational duration with PGS as covariate (with or without the batch and clinical covariates described above). For comparison, we reran this model, but excluding all variants further than 50 kb away from any of the top maternal hits (weights were unchanged for the retained variants). We also refitted the PAMM model for each variant with PGS as an additional constant-effect covariate.

The PGS was then categorised into quintiles and a PAMM fitted with a smooth term for each category (except the middle one as the reference). This was repeated with tertile or septile categories of PGS instead. Genotyping batch and the clinical covariates were kept as constant-effect covariates. We also show the risks of preterm and very preterm delivery (fraction of spontaneous births in this sample <259 or <224 days of gestation, respectively) and their confidence intervals obtained with R *prop*.*test*.

In the exploratory analyses, we first inspected the total per-individual counts of rare deleterious alleles, which we defined as having a negative effect of at least 0.001 days in the PGS and frequency <10%. We similarly plotted the counts of deleterious recessive genotypes – minor homozygous genotypes for alleles with < –0.001 days effect in the PGS. We then ran a principal component (PC) analysis on the maternal genetic data, using only autosomal PGS variants, pruned to reduce dependency with PLINK’s *--indep-pairwise 100 10 0*.*2* command. Top 10 PCs were extracted. All PCs and the two counts were tested for association with preterm or very preterm delivery in logistic regression models, adjusting for the PGS and genotyping batch. Variant loadings on PC3 were taken as reported by PLINK, and the Genome Reference Consortium boundaries of the MHC region were retrieved from https://www.ncbi.nlm.nih.gov/grc/human/regions/MHC?asm=GRCh37.

## Data Availability

Summary GWAS statistics produced in the present work will be made available in the GWAS Catalogue upon manuscript acceptance. Individual-level data from the MoBa cohort is available for researchers by application through helsedata.no. 23andMe, Inc. data used for training of the polygenic score is available through 23andMe to qualified researchers under an agreement with 23andMe (see research.23andme.com). Code used for the analyses is available at https://github.com/PerinatalLab/time-varying.

## Data availability

All code for replicating the analyses presented here is available at https://github.com/PerinatalLab/time-varying. Summary statistics from the GWAS performed as part of this project (foetal and maternal GWAS of gestational duration and preterm delivery) are available from the MoBa website (*link available after acceptance for publication*) and the GWAS Catalogue (*link available after acceptance for publication*). The full GWAS summary statistics for the 23andMe data used in the construction of PGS are available through 23andMe to qualified researchers under an agreement with 23andMe that protects the privacy of the 23andMe participants. Please visit https://research.23andme.com/collaborate/#dataset-access/ for more information and to apply to access the data. A simplified set of weights for the polygenic score of gestational duration, trained excluding 23andMe, Inc. data, is available in the PGS Catalog.

The consent given by the participants does not permit public access of data at the individual level. Researchers can apply for access to data through helsedata.no. Access requires approval from the Regional Committees for Medical and Health Research Ethics in Norway and an agreement with MoBa.

## Acknowledgements

The Norwegian Mother, Father and Child Cohort Study is supported by the Norwegian Ministry of Health and Care Services and the Ministry of Education and Research. We are grateful to all the participating families in Norway who take part in this on-going cohort study.

We thank the Norwegian Institute of Public Health (NIPH) for generating high-quality genomic data. This research is part of the HARVEST collaboration, supported by the Research Council of Norway (#229624). We also thank the NORMENT Centre for providing genotype data, funded by the Research Council of Norway (#223273), South East Norway Health Authorities and Stiftelsen Kristian Gerhard Jebsen. We further thank the Center for Diabetes Research, the University of Bergen for providing genotype data and performing quality control and imputation of the data funded by the ERC AdG project SELECTionPREDISPOSED, Stiftelsen Kristian Gerhard Jebsen, Trond Mohn Foundation, the Research Council of Norway, the Novo Nordisk Foundation, the University of Bergen, and the Western Norway Health Authorities. We would like to thank the research participants and employees of 23andMe for making this work possible.

B.J. received funding from The Swedish Research Council, Stockholm, Sweden (2019-01004), The Research Council of Norway, Oslo, Norway (FRIMEDBIO #547711), March of Dimes (#21-FY16-121), Agreement concerning research and education of doctors (ALFGBG-965353). J.J. received funding from Lilla Barnets Foundation. Research by B.J. was also supported by the Eunice Kennedy Shriver National Institute Of Child Health & Human Development of the National Institutes of Health under Award Number R01HD101669. The content is solely the responsibility of the authors and does not necessarily represent the official views of the National Institutes of Health.

