## Supplementary Material for "Time-varying effects are common in genetic control of gestational duration"

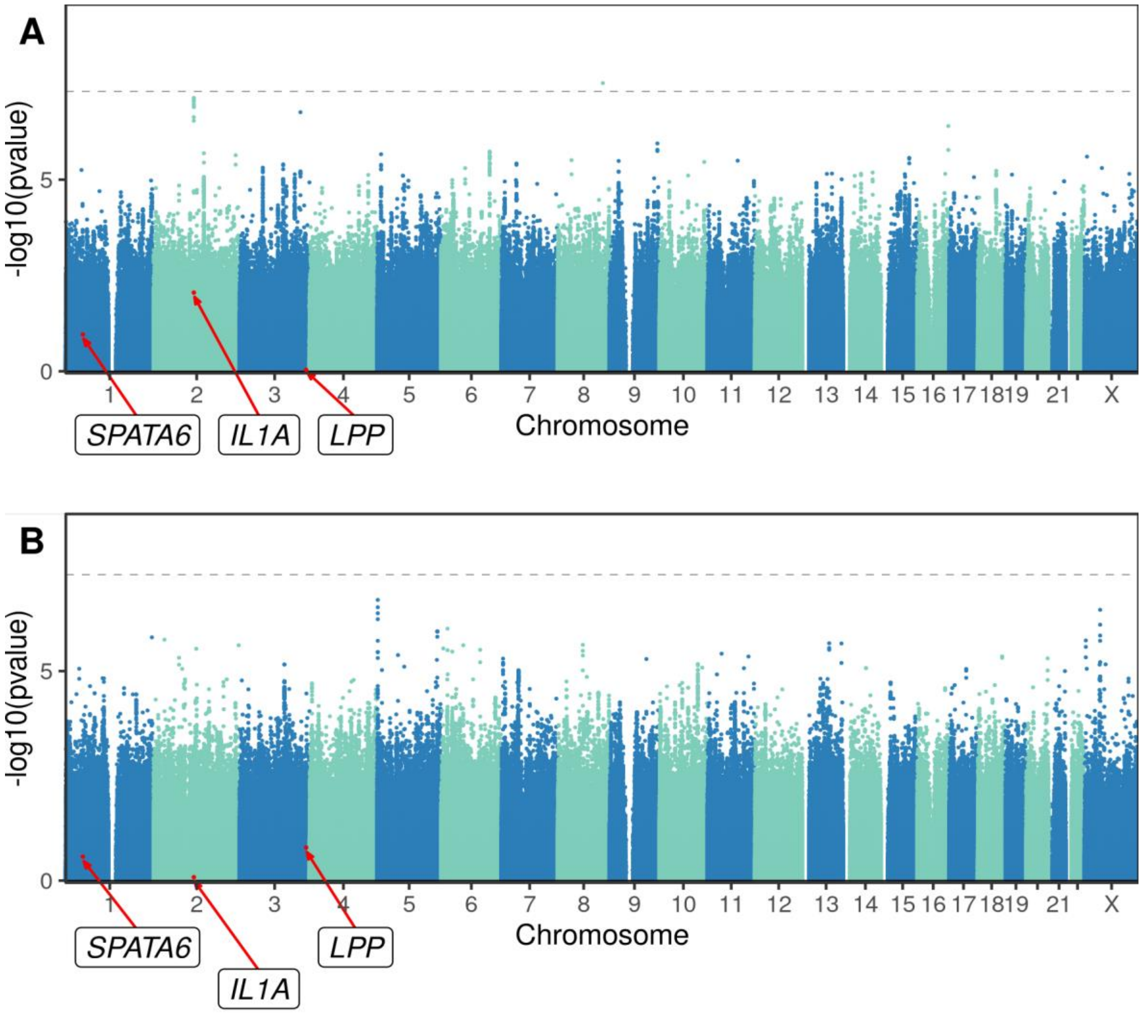


**Figure S1.** Manhattan plot illustrating the foetal GWAS results for gestational duration (A) and preterm delivery (B). The dashed line marks the significance threshold. Note the one genome-wide significant association for gestational duration in chromosome 8. No genome-wide significant association in the foetal GWAS of preterm delivery was detected. The variants marked in red, and the corresponding loci, were reported as significant in the largest previous foetal GWAS meta-analysis of gestational timing (Liu et al. 2019).

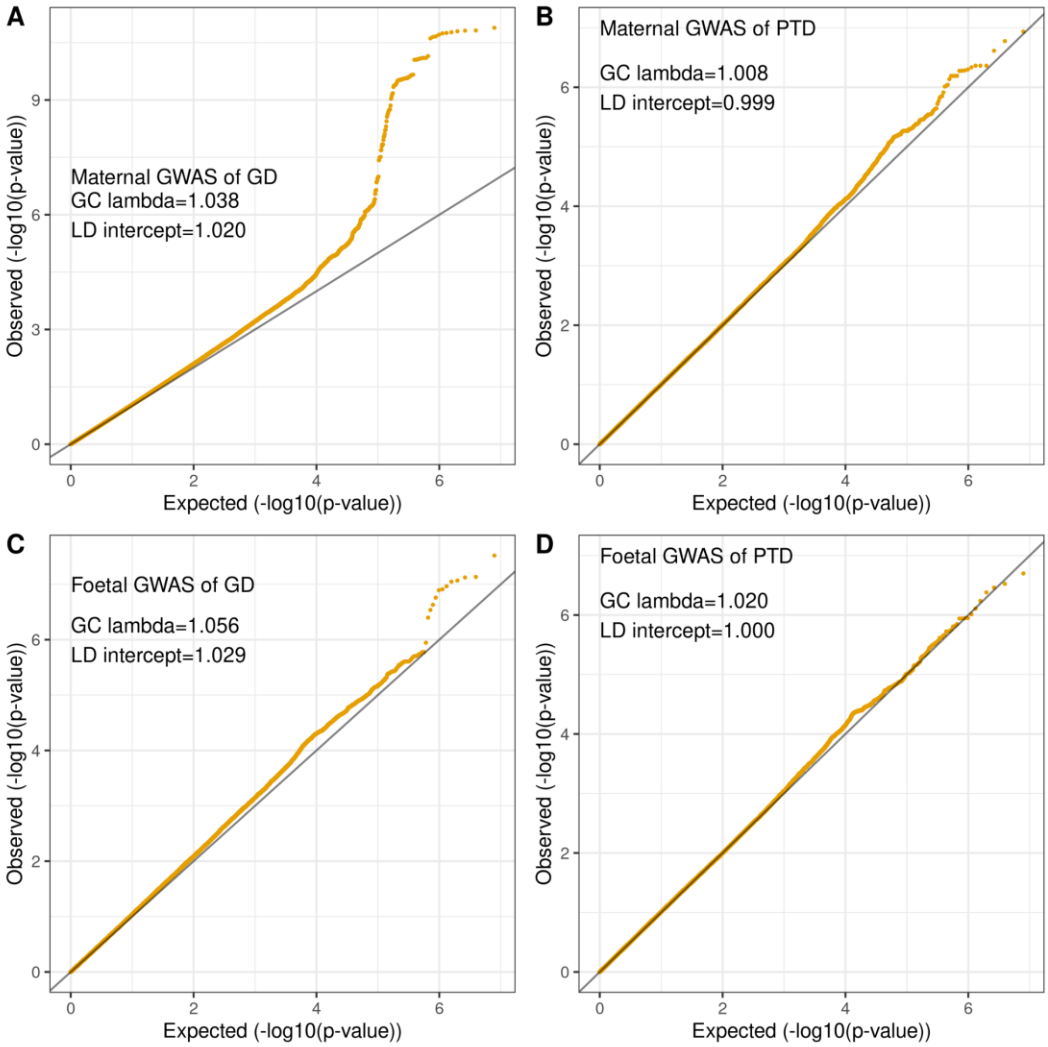


**Figure S2**. Quantile-quantile plots of maternal (A,B) or foetal (C,D) GWASs of gestational duration (A,C) and preterm delivery (B,D). Lambda genomic control and LD score regression intercept values are shown in the labels, and show little to none test statistic inflation in all cases.

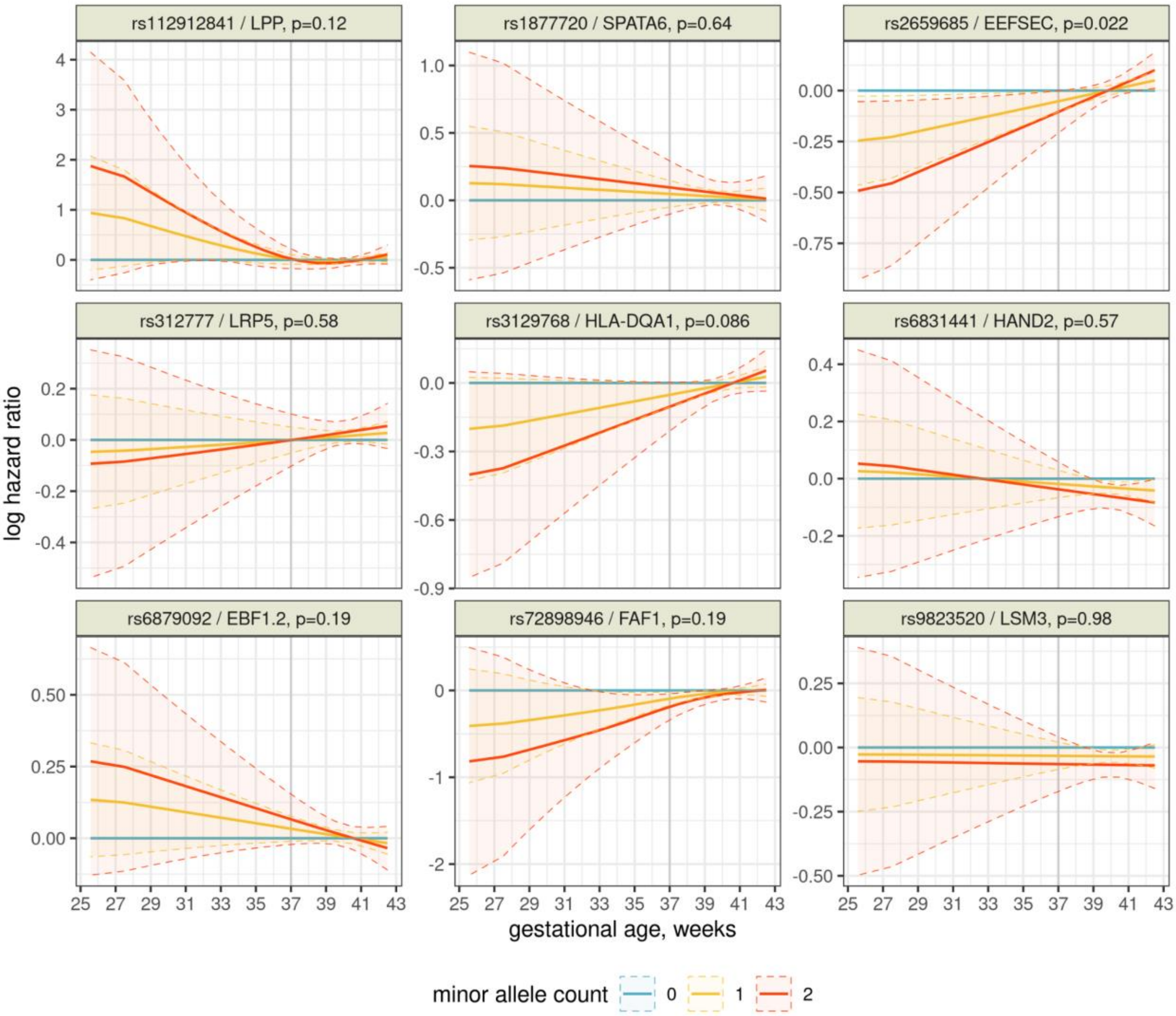


**Figure S3**. Effects of 9 genetic variants on delivery timing throughout gestation, estimated by a flexible PAMM. Variants shown here were detected in a previous meta-analysis, but had no significant effects in the GWAS in this cohort. For each week of gestation, the line is the estimated instantaneous log hazard ratio (positive=risk of birth is currently increased) and its 95 % confidence interval is shaded. *EBF1.2* is a second association reported near the *EBF1* gene. Note that the minor allele count was used as a continuous covariate in the analysis, so the effects of 1 and 2 minor alleles are constrained to be proportional.

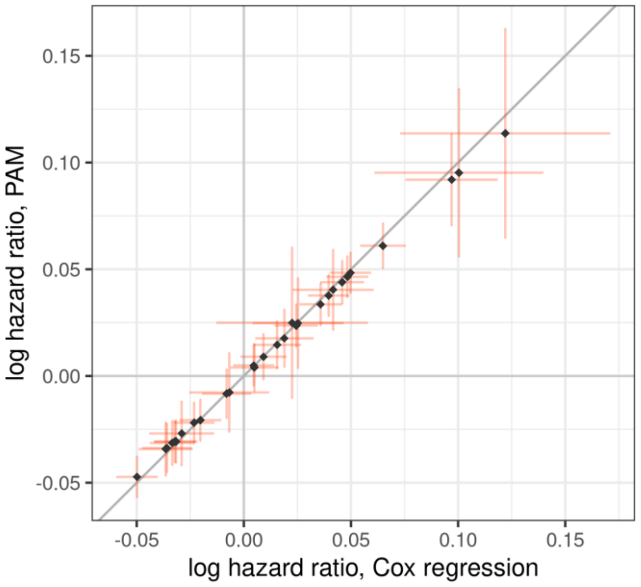


**Figure S4**. Comparison between the variant effect estimates from Cox regression and a simplified PAM with only constant (parametric) effects. Points are the estimates and bars are +/- 1 SE. The diagonal grid line representing equal effects is highlighted.

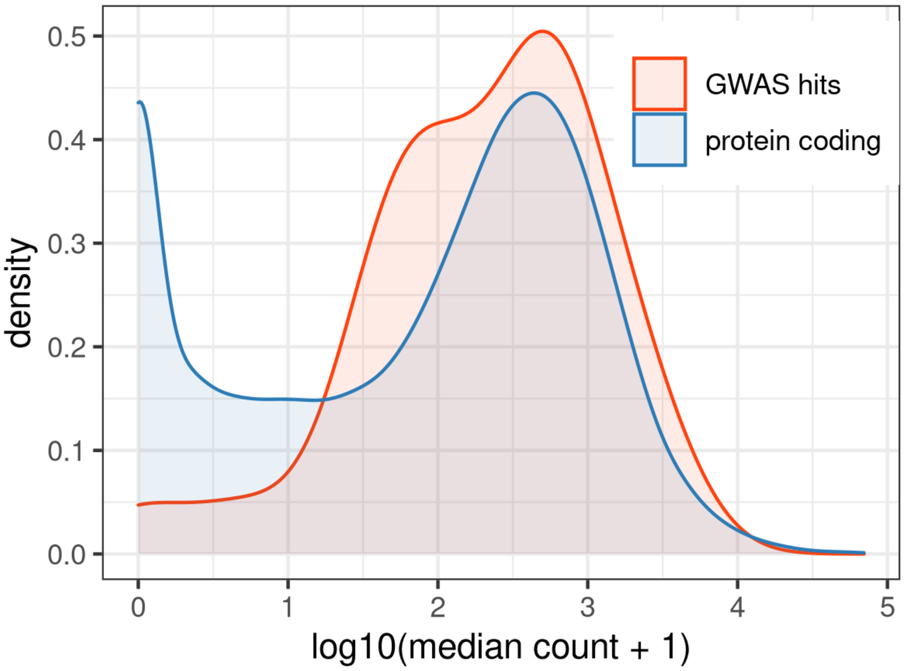


**Figure S5**. Distribution of median read counts for different genes in placental expression data (median taken over samples). “GWAS hits” are the 28 distinct genes corresponding to the loci analysed in detail in this study, “protein coding” are all other protein coding genes identified in the Ensembl database.

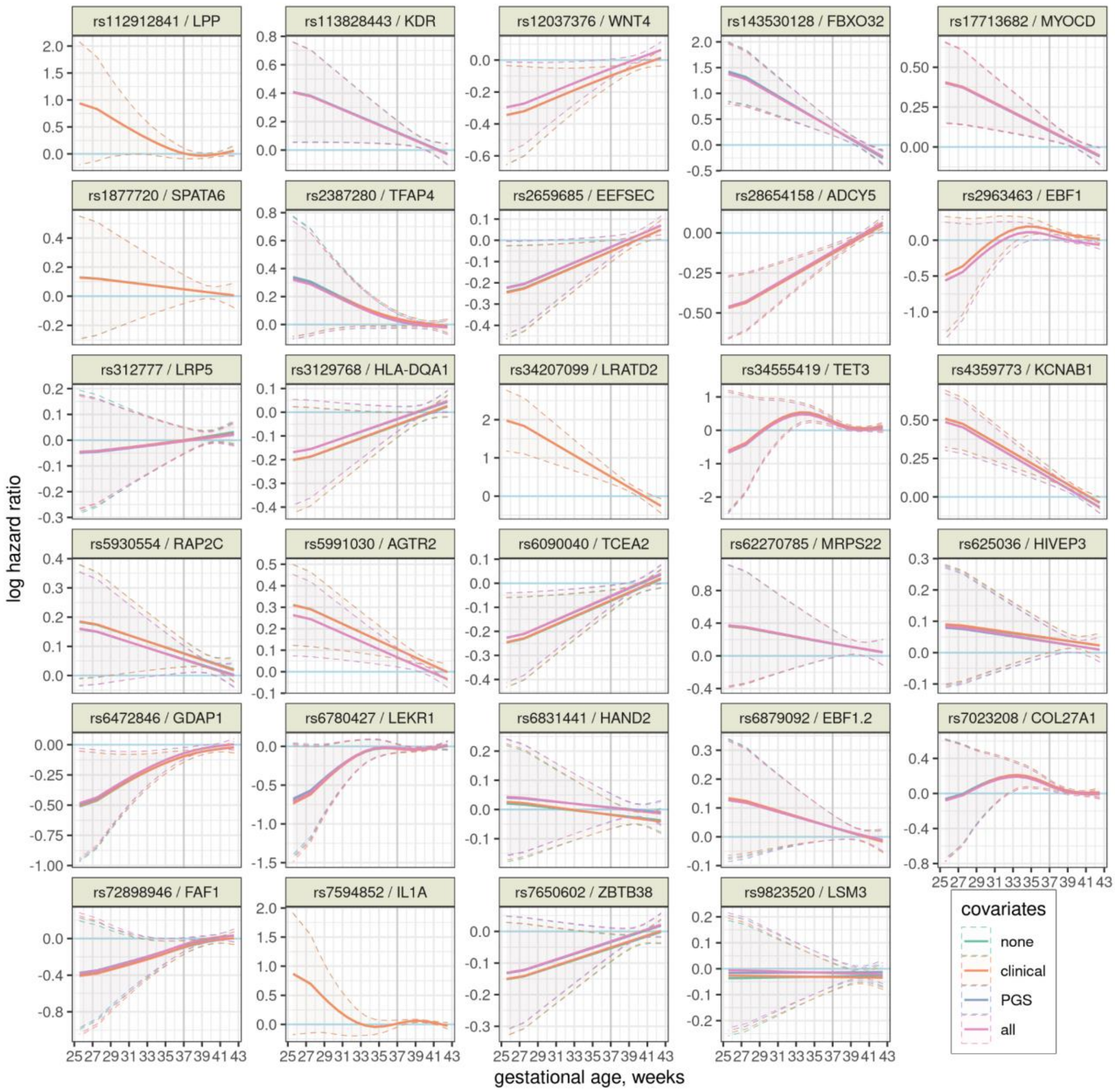


**Figure S6**. Effects of 29 genetic variants on delivery timing throughout gestation, estimated by flexible PAMMs with different covariates. The effects are shown per one copy of minor allele. For each week of gestation, the line is the estimated instantaneous log hazard ratio (positive=risk of birth is currently increased) and its 95 % confidence interval is shaded. PGS: maternal polygenic score, only shown for the maternal loci. Note that the lines with or without clinical covariates overlap almost perfectly.

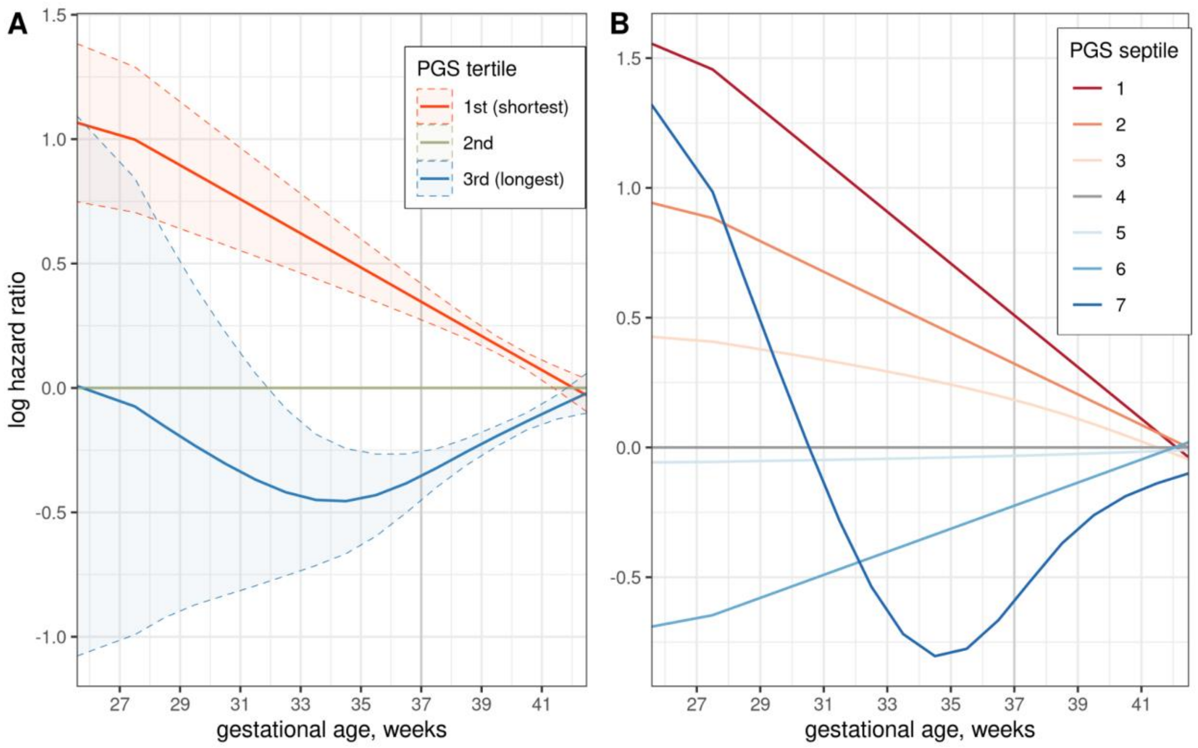


**Figure S7.** Effects of the gestational duration polygenic score throughout gestation, estimated by a flexible PAMM. The polygenic score is split into 3 (A) or 7 (B) categories. For each week of gestation, the line is the estimated instantaneous log hazard ratio (positive=risk of birth is currently increased). The 95 % confidence intervals are shaded in the 3-group plot, but for clarity not shown in the 7-group plot.

| locus | rsid | chr | pos | ref | eff | EAF | lin.beta | pc.sm.p | pc.int.p | cox.sch.p |
|---|---|---|---|---|---|---|---|---|---|---|
| KCNAB1 | rs4359773 | 3 | 155862524 | A | G | 0.43 | -0.65 | 1.2e-06 | 2.7e-07 | 7.1e-05 |
| FBXO32 | rs143530128 | 8 | 124603784 | G | A | 0.02 | -1.5 | 4.1e-06 | 2.5e-06 | 0.00039 |
| LRATD2 | rs34207099 | 8 | 127494476 | A | T | 0.014 | -2.7 | 4.3e-06 | 3.1e-06 | 0.00036 |
| ADCY5 | rs28654158 | 3 | 123112292 | T | C | 0.38 | 0.52 | 7.3e-06 | 4.6e-06 | 1.1e-05 |
| MYOCD | rs17713682 | 17 | 12416363 | G | T | 0.19 | -0.4 | 0.0023 | 0.0019 | 0.011 |
| TET3 | rs34555419 | 2 | 74235926 | A | G | 0.055 | -1 | 0.0033 | 0.015 | 0.097 |
| AGTR2 | rs5991030 | X | 115129904 | T | C | 0.43 | -0.48 | 0.0053 | 0.0043 | 0.017 |
| EBF1 | rs2963463 | 5 | 157895049 | C | T | 0.27 | -0.55 | 0.011 | 0.01 | 0.0091 |
| TCEA2 | rs6090040 | 20 | 62692060 | A | C | 0.46 | 0.28 | 0.016 | 0.014 | 0.0058 |
| COL27A1 | rs7023208 | 9 | 116935764 | C | G | 0.34 | -0.33 | 0.019 | 0.015 | 0.12 |
| EEFSEC | rs2659685 | 3 | 128122396 | G | A | 0.24 | 0.2 | 0.022 | 0.019 | 0.0067 |
| GDAP1 | rs6472846 | 8 | 75315146 | G | C | 0.41 | 0.48 | 0.035 | 0.017 | 0.095 |
| KDR | rs113828443 | 4 | 55895282 | C | T | 0.075 | -0.42 | 0.039 | 0.034 | 0.07 |
| IL1A | rs7594852 | 2 | 113521754 | C | T | 0.46 | -0.24 | 0.042 | 0.29 | 0.15 |
| WNT4 | rs12037376 | 1 | 22462111 | G | A | 0.17 | 0.36 | 0.043 | 0.021 | 0.084 |
| HLA-DQA1 | rs3129768 | 6 | 32595083 | T | G | 0.22 | 0.19 | 0.086 | 0.074 | 0.071 |
| LEKR1 | rs6780427 | 3 | 156697097 | A | G | 0.4 | 0.33 | 0.11 | 0.14 | 0.051 |
| LPP | rs112912841 | 3 | 187987683 | A | G | 0.073 | 0.021 | 0.12 | 0.93 | 0.32 |
| RAP2C | rs5930554 | X | 131312089 | T | C | 0.32 | -0.37 | 0.15 | 0.14 | 0.23 |
| ZBTB38 | rs7650602 | 3 | 141147414 | T | C | 0.47 | 0.26 | 0.16 | 0.15 | 0.18 |

| | | | | | | | | | | |
|---|---|---|---|---|---|---|---|---|---|---|
| TFAP4 | rs2387280 | 16 | 4339684 | A | T | 0.27 | -0.22 | 0.18 | 0.12 | 0.22 |
| EBF1.2 | rs6879092 | 5 | 158058432 | T | G | 0.5 | -0.15 | 0.19 | 0.17 | 0.3 |
| FAF1 | rs72898946 | 1 | 50959262 | A | C | 0.11 | 0.28 | 0.19 | 0.072 | 0.42 |
| MRPS22 | rs62270785 | 3 | 139004333 | A | G | 0.016 | -0.79 | 0.49 | 0.48 | 0.78 |
| HIVEP3 | rs625036 | 1 | 42270779 | G | A | 0.36 | -0.29 | 0.55 | 0.54 | 0.82 |
| HAND2 | rs6831441 | 4 | 174733296 | T | G | 0.31 | 0.13 | 0.57 | 0.56 | 0.99 |
| LRP5 | rs312777 | 11 | 68107264 | C | G | 0.26 | -0.034 | 0.58 | 0.52 | 0.62 |
| SPATA6 | rs1877720 | 1 | 48824407 | C | T | 0.054 | -0.32 | 0.64 | 0.62 | 0.48 |
| LSM3 | rs9823520 | 3 | 14293832 | G | A | 0.22 | 0.21 | 0.98 | 0.95 | 0.88 |

**Table S1**. The top variants tested in time-varying models, and comparison of three methods for testing if the effects vary in time. Ref, eff – reference and effect alleles; EAF – effect allele frequency; lin.beta – effect size on gestational duration in days, from linear regression; pc.sm.p, pc.int.p, cox.sch.p are p-values from, respectively, a PAM model with smoothly varying effects, a PAM model with a linear interaction term, and a Schoenfeld residuals test with a Kaplan-Meier transform. Note that *EBF1.2* is used throughout this paper to refer to a second association in the *EBF1* locus.
